# Four Models of Wastewater-Based Surveillance for SARS-CoV-2 in Jail Settings: How Monitoring Wastewater Complements Individual Screening

**DOI:** 10.1101/2023.08.04.23293152

**Authors:** S Kennedy, AC Spaulding, SWANSS writing group *

**Affiliations:** Emory University

## Abstract

**Objective:** To describe four unique models of implementing Wastewater Based Surveillance (WBS) for SARS-CoV-2 in jails of graduated sizes and differing architectural designs.

**Methods:** This study summarizes how jails of Cook County (Illinois, average daily population [ADP] 6000), Fulton County (Georgia, ADP 3000, Washington DC (ADP 1600) and Middlesex County (Massachusetts, ADP 875) initiated WBS between 2020 and 2023.

**Results:** Positive signal for SARS-CoV-2 via WBS can herald new onset of infection in a previously uninfected housing unit of a jail. Challenges in implementing WBS included political will and realized value, funding, understanding of the building architecture, and the need for granularity in the findings.

**Conclusions:** WBS has been effective for detecting outbreaks of SARS-CoV-2 in differing sized jails, both those with dorm-based and cell-based architectural design.

**Policy implications:** Given its effectiveness in monitoring SARS-CoV-2, WBS provides a model for population-based surveillance in carceral facilities for future infectious disease outbreaks.

## Introduction

SARS-CoV-2 transmission rates spiked early in carceral populations^1^ due to congregate living, poor ventilation, and abrupt population shifts in cellblocks, which facilitate transmission of airborne pathogens.^2–4^ Pre-pandemic, the US led the world in incarceration^5^ and its jails (short-term correctional facilities) averaged 11 million admissions yearly,^6^ representing 7-8 million individuals, when accounting for repeat admissions.^7^ As of January 2023, COVID-19 had caused 3,181 deaths in US prisons and jails.^8^ Surveillance and interventions to decrease mortality in custody populations are crucial.

Centers for Disease Control and Prevention (CDC) guidance on COVID-19 Prevention Outbreak Management in Carceral Settings focused on individual testing (e.g., nasal swabs), quarantine, isolation, and mitigation. Recent revisions add Wastewater Based Surveillance (WBS) as another mitigation strategy. WBS consists of testing wastewater to identify pathogens which can then be linked to a population source. This mass testing strategy can then be supplemented by individual diagnostic tests to identify source cases.

Initially developed 70 years ago for surveilling pathogens shed in feces,^9–11^ WBS has proven most useful with diseases that spread rapidly, have non-specific symptoms, and are often asymptomatic.^12^ These characteristics align with COVID-19. Because SARS-CoV-2 persists in fecal matter, WBS represents a practical and effective means of mass surveillance. Several college dormitory studies provided evidence that WBS is a sensitive, low-cost, non-invasive surveillance tool to detect early COVID-19 at an institutional level.^13-^19 Nonetheless, no guidance for operationalizing WBS in jails exists. We conducted qualitative studies exploring acceptability of WBS before introducing it at Fulton County Jail in Atlanta.^20, 21^ Given the need for early and affordable outbreak detection in congregate settings, implementation of WBS could potentially save time, preserve resources, and improve health. Here we describe how four jails attempted to implement WBS for early detection and management of SARS-CoV-2.

## Methods

### Setting

Four jail systems participated in this study: Cook County Jail (CCJ, Chicago, Illinois); Fulton County Jail (FCJ, Atlanta, Georgia); Middlesex House of Corrections and Jail (MHOCJ, North Billerica, Massachusetts); and the District of Columbia Jail (DCJ, Washington, DC). Each secured funding early in the pandemic to be early adopters of WBS and were willing to engage in an implementation study to sustain the surveillance. This study includes jails of differing sizes, architectural designs, sewer system configurations, and funding levels.

### Wastewater Collection and Laboratory Methods

Two dominant methods of gathering wastewater samples exist. Grab samples are a snapshot of viral levels. Longitudinal measurement of viral levels can be derived using Moore swabs, 4” × 4” gauze squares tied in the middle with a fishing line (Figure 1) and dropped in sewer lines for 24-48 hours. By collecting either grab or Moore samples, WBS monitors the presence and concentration of specific gene sequences.^22^ These two collection methods are analogous to a fingerstick blood glucose (single point measurement) and a hemoglobin A1C (period measurement) for monitoring blood glucose in diabetic patients. Community-level surveillance can then commence wastewater sampling and obtainment of a laboratory based semi-quantitative measurement of virus in the sample.^23^ Results are presented as Cycle Thresholds (Ct), where the signal is no longer visible. This semi-quantitative measurement provides a rough comparison of relative levels of virus over time.

**Figure 1.**
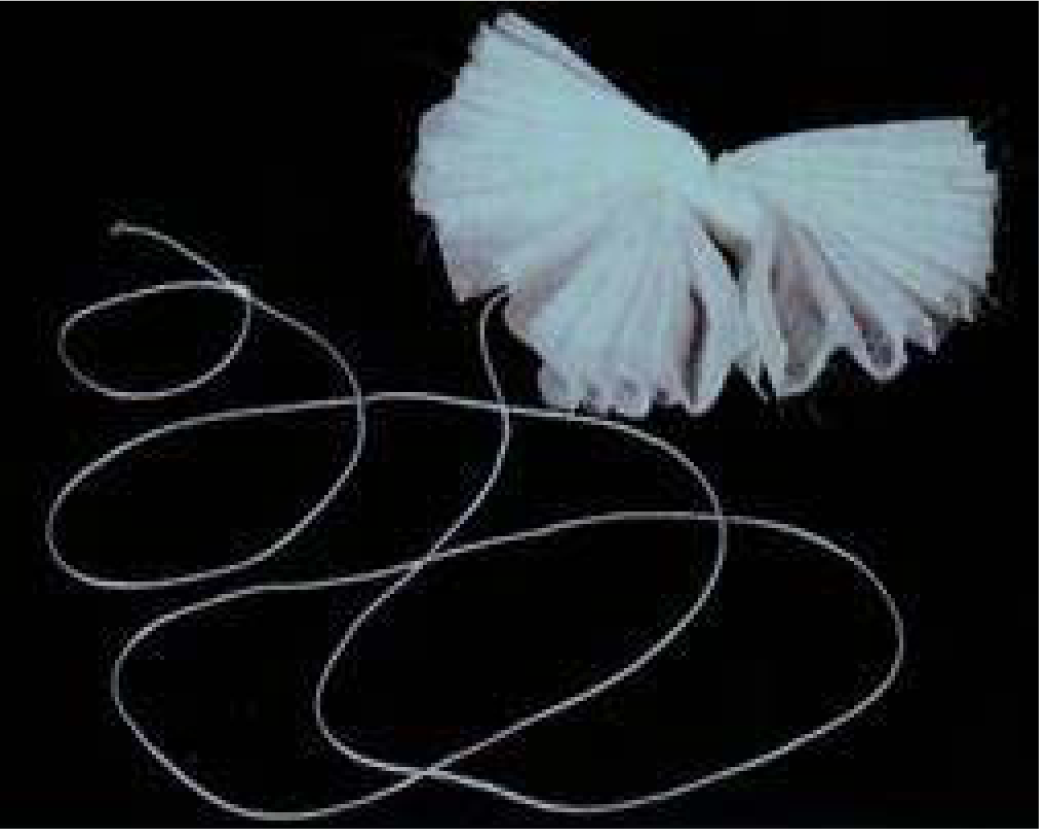
Moore Swab. A 4” by 4” gauze square, tied together with fishing line, which is suspended in wastewater for 24 hours.

### Data Collection

Each active site had one or more authors participating in wastewater surveillance. For each site, we explored and compared initial funding, starting points, physical structure of the jails, process for wastewater sample collection, and laboratory methods.

## Results

### WBS Funding, Starting Points and Laboratory Linkage

Each jail included in this analysis established external partners that provided an established laboratory for wastewater testing. In Chicago, Illinois, a city-wide project monitoring wastewater for SARS-CoV-2 started in the fall of 2020 and included the 16 buildings of the <u>CCJ</u> system. The Walder Foundation funded the Chicago Prototype Coronavirus Assessment Network Node (PCANN). Specimens were processed at the University of Illinois at Chicago.

<u>FCJ</u> in Atlanta, Georgia, had WBS externally funded by the NIH through Ceres Nanoscience (Manassas, Virginia). The company subcontracted with Emory University in 2021 to pilot testing wastewater at points around the city of Atlanta. This included using a Nanotrap particle-based viral concentration method developed by Ceres Nanoscience, through a KingFisher Apex robotic platform (Thermo Fisher Scientific, USA). Additionally, Emory University received a grant from the Bill & Melinda Gates Foundation to demonstrate the efficiency of self-collected nasal swabbing of jail residents.

<u>MHOCJ</u> is the smallest jail included in this report although ranked as a medium size jail by national standards.^24^ Funding and laboratory resources were provided by the Sherriff’s Department and Biobot Analytics (Cambridge, MA), respectively. MHOCJ began weekly sampling of wastewater in April 2021.

<u>DCJ</u> had its WBS established by the Washington DC Department of Health (DOH), recipient of a grant in 2020 from the CDC’s National Wastewater Surveillance System which included the jail. Surveillance began at school sites before the jail then was temporarily placed on hold due to challenges in meeting the site collection criteria for proper testing and sampling. Currently, DOH contracts with EA Engineering for sample collection and The DC Public Health Lab processes the specimens. Routine sampling at the DCJ began in March 2023. The data collected has been shared with the jail since June 2023, and will be used to mitigate future outbreaks. Long term surveillance is expected to continue at DCJ.

### WBS Operations and Jail Structure

<u>CCJ</u>: Researchers at the University of Illinois at Chicago School of Public Health tested wastewater samples as part of a larger COVID-19 surveillance project with the Discovery Partners Institute.^25^ Testing involved collection of grab samples and Moore swabs. Over 30 possible collection sites were identified during the initial site visit to the multi-building CCJ complex (Figure 2a). Monitored sites were selected weekly by the infection control lead (author CJZ). Initially, six sites were tested twice weekly, with periodic adjustments based on disease activity. Collection site selection was initially based on confirmed clinical activity or suspicion of new infections to validate the connection between wastewater readings and cases. Once this link was established, the strategy shifted to focus on the areas where suspicion of infection was low, as an early warning of new outbreaks. For buildings in which known cases were rare and the wastewater remained negative, active surveillance of individuals was minimized.

**Figure 2.**
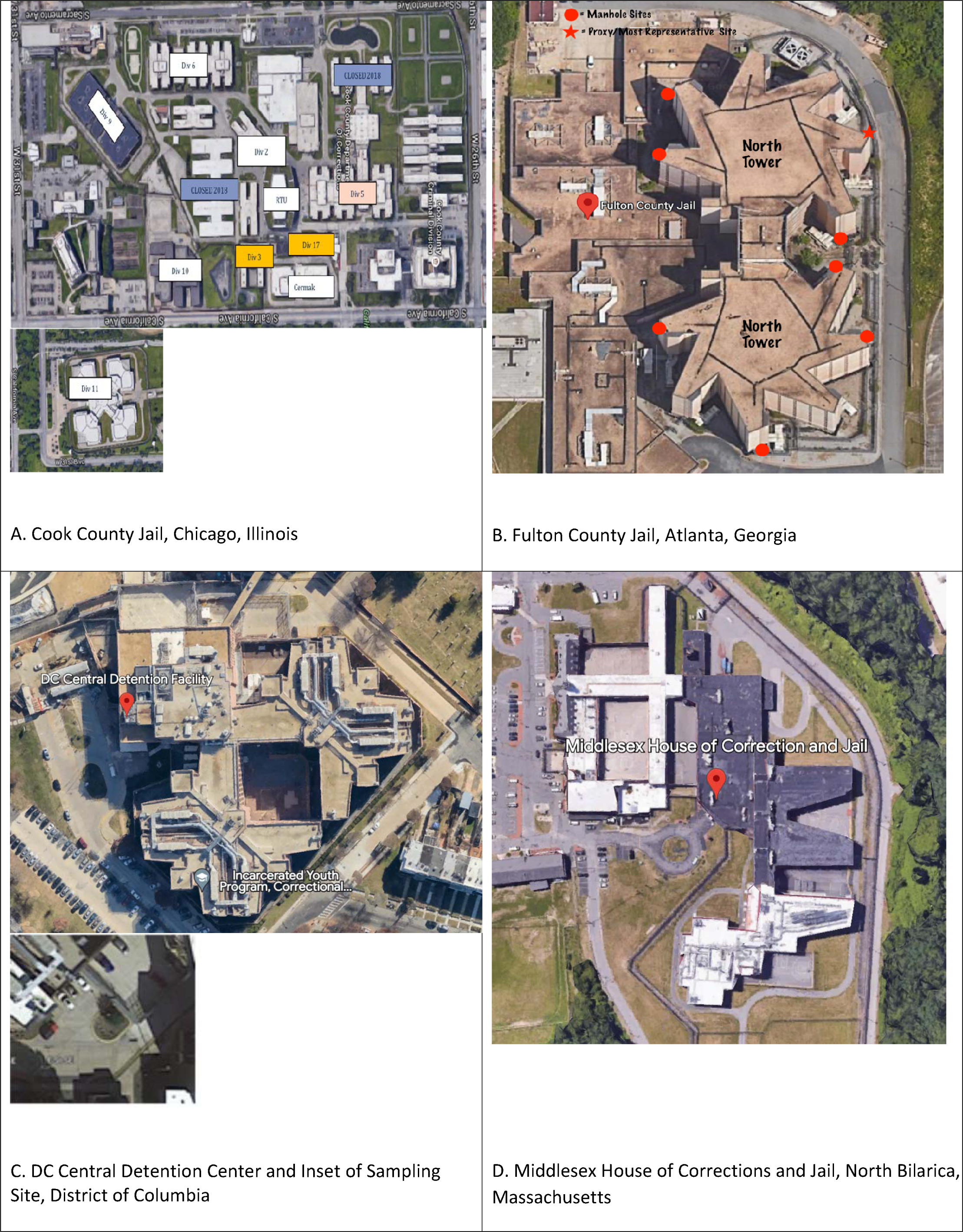
Aerial photographs of jails of (a) Cook County, Illinois, (b) Fulton Count, GA (c) Middlesex County, MA and (d) DC Ja Map with location of housing units and manhole points. Source: Google Maps.

<u>FCJ</u>: In April of 2021, the Emory University Center for Global Safe WASH piloted wastewater collection at the jail. Its Environmental Microbiology laboratory performed real-time, quantitative reverse transcription-polymerase chain reaction (RT-qPCR), viral concentration, and RNA extraction on the specimens. The Emory team employed technology similar to that used in university dorm testing.^22, 26^ Starting in June 2021, weekly water samples were collected. With a jail-representative escort, Emory staff collected Moore swabs and grab sampling of 40 mL of wastewater weekly from 6 out of 11 accessible manholes on the property (Figure 2b). The semi-quantitative results were reported the following day.^27^ The jail’s medical team deliberated with custody staff regarding individual testing and isolation. The main FCJ complex consists of a 7-floor structure with two towers; each tower floor contains six housing units (Textbox 1 and Figure 3b). At the start of the study, FCJ’s sewage system blueprints could not be located. Tracer dye (EcoClean Solutions, Copiague, NY) was poured into the plumbing system at various source locations, with study staff stationed at manholes relaying which color appeared when. Plumbing blueprints were located in October 2022. By early 2023, the study team validated the flow of sewage lines shown in the blueprint to determine the precise location of the wastewater source. (Textbox 1). With future outbreaks of pathogens that can be tracked in wastewater, the precise location can thus be narrowed down to one of six specific housing units.

**Figure 3.**
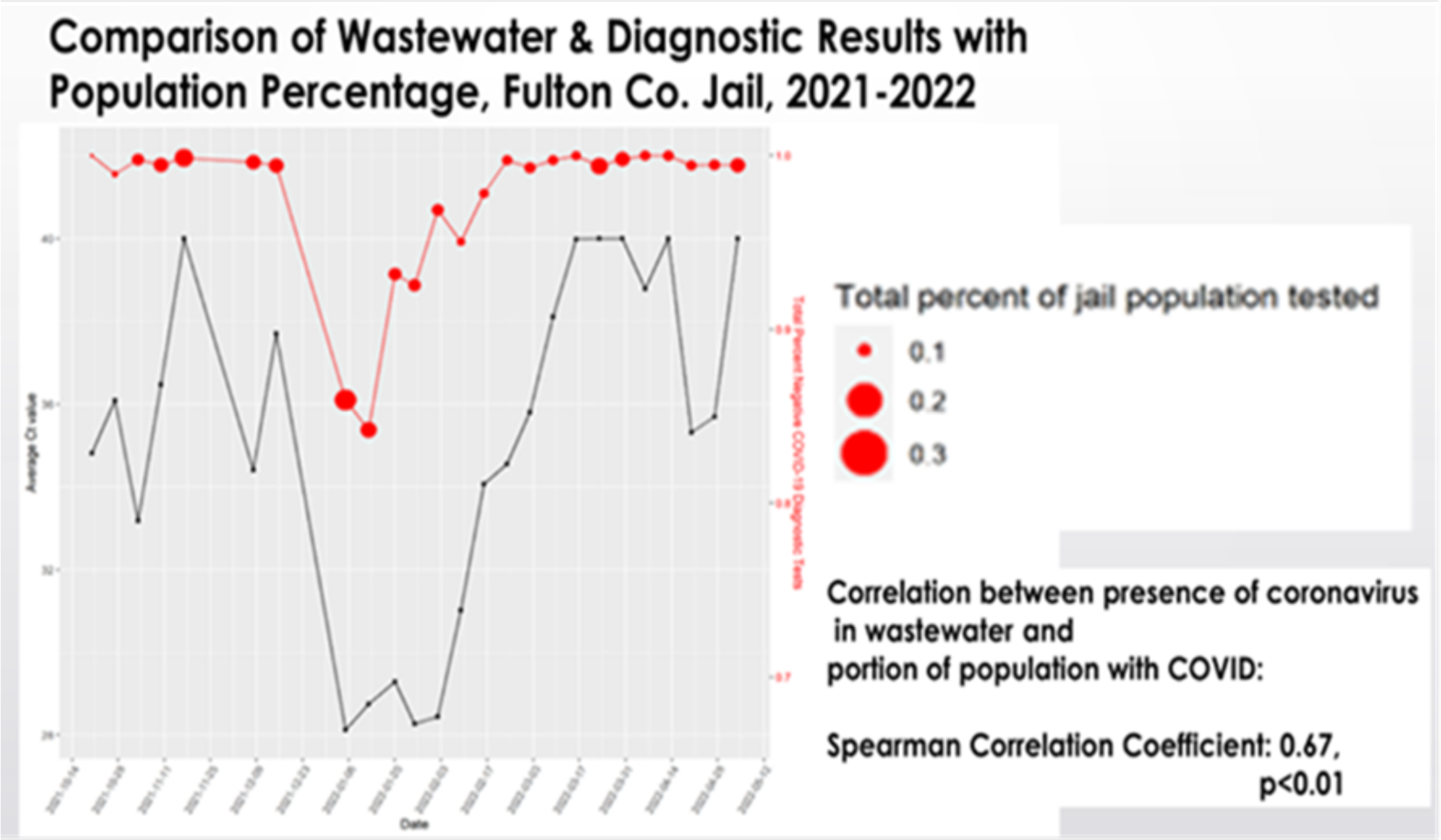
Source: Saber et al. Wastewater Surveillance for SARS-CoV-2 in an Atlanta, Georgia Jail: A study of the feasibility of wastewater monitoring and correlation of building wastewater and individual testing results medRxiv 2023.05.17.23290000; doi:https://doi.org/10.1101/2023.05.17.23290000

<u>MHOCJ</u> began wastewater testing in April 2021, every Monday. Wastewater was sampled from the single manhole site where sewage exited the jail campus using an automated sampler^28^ (Figure 2c). The specimen was analyzed by a commercial laboratory, Biobot Analytics (Cambridge, MA). Results were delivered electronically to the sheriff’s office 48 hours later. Increased viral concentrations in the wastewater prompted meetings with the infectious disease consultant (author AGW) and jail staff to discuss enhanced individual testing and mitigation.

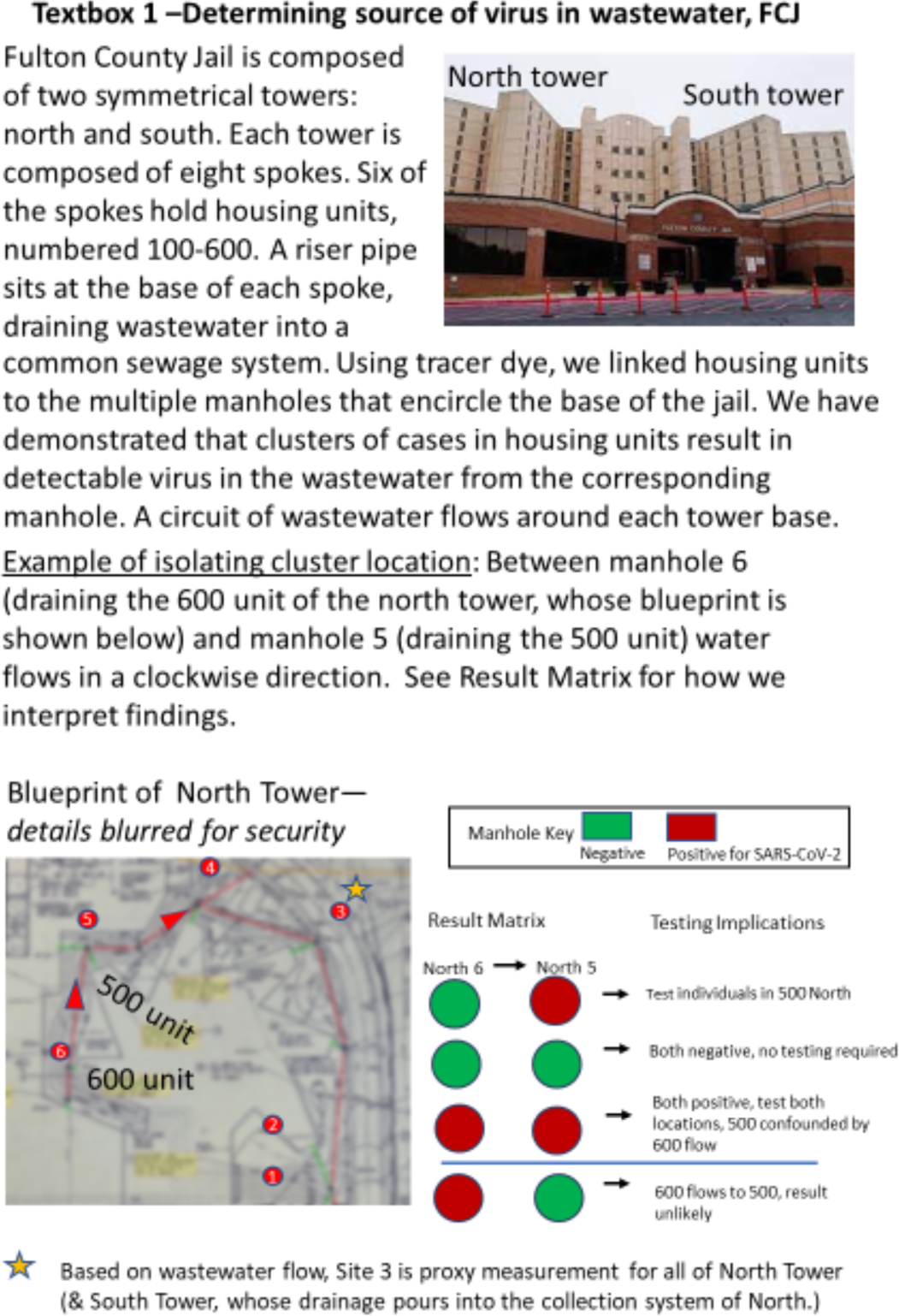

<u>DC Jail</u>: The DC DOH is using an automatic sampler and selected a single site for collection near the infirmary and a housing unit for mid-to-long-term residents (Figure 2d). Conversations are underway between the DC DOH and the Department of Corrections medical services about how clinical services can use the data generated.

### Individual Testing Procedures

Each of the four jails developed dynamic COVID-19 response protocols that included resident and staff testing. Three have incorporated WBS in their operations.

<u>CCJ</u> used a Rapid PCR (IDNow, Abbot Laboratories, Abbot Park III) test at intake and in the urgent care unit. For screening of housing units after exposure, before medical procedures, or prior to prison transfer, staff collected swabs for laboratory-based PCR testing performed at the John H. Stroger Jr. Hospital (Chicago, IL). Prior to the emergence of the Omicron variant, entrants were quarantined in intake housing, and PCR testing was used to clear them for transfer to the general population. This procedure was revised after CDC guidelines changed.

<u>FCJ</u> conducted opt-out rapid testing of individuals at intake as well as point-of-care testing for suspected cases in population using point-of-care antigen tests (BinaxNow, Abbot Laboratories, Chicago IL, through January 2022; QuickVue, QuidelOrtho, San Diego CA, from February 2022 onwards). Periodic mass testing was performed by a team from Rollins School of Public Health. The Gates Foundation suggested random population testing be done via self-collected SteriPack nasal swabs [SteriPack USA, Lakeland FL], collection devices for molecular diagnostic testing (see Supplement, Section 3,). Prior to piloting this strategy, the Emory team held three focus groups of recently released individuals from local jails to gauge acceptability; these groups specifically endorsed this collection strategy.20 Once mass testing began, the Emory team tracked the relationship between COVID-19 diagnostic test positivity rates and levels of SARS-CoV-2 in wastewater.

Specimens collected at FCJ were tested by Northwell Health Laboratory who used an LGC, Biosearch Technologies [Middlesex, UK] SARS-CoV-2 ultra-high-throughput End-Point RT-PCR Test (BT-SCV2-UHTP-EP) with 100% sensitivity. Jail medical staff could access results the next day. The relationship between COVID-19 individual test positivity rates with levels of SARS-CoV-2 in the wastewater was found to be correlated (Figure 3a).29 In early 2022, the community prevalence of COVID-19 spiked. At that time, if custody staff opted out of taking a vaccine, they had to provide negative individual PCR tests twice weekly. Emory extended access to the Gates-funded tests at custody’s roll calls and could see the officers’ test positivity rate was likewise high.

SARS-CoV-2 individual testing at <u>MHOCJ</u> started with PCR testing in April 2020. In November 2021, PCR testing was replaced by rapid antigen testing. <u>DC DOC</u> procedures for screening are described in a previous publication.^30^

### Using WBS and Individual Testing to Inform Mitigation Strategies

Targeted testing can follow a newly positive wastewater signal. The number of possible wastewater collection points, shown in Table 1, is proportional to the size of each jail. With one collection point, the readings are akin to a community viral load. However, when wastewater can be collected from multiple collection sites and the source is known to originate from a particular location in the jail, one or more positive signals informed the jail to consider testing where there may be highest risk of infection/spread. When positive sites were widespread, negative sites signaled where cases were not occurring and, therefore, resources could be spared. CCJ demonstrated the most progress in utilizing WBS to locate and respond to outbreaks identified by WBS at various jail housing units. Once the Omicron variant of SARS-CoV-2 was widespread in the community at-large, wastewater was consistently positive in most buildings. This guided the jail to focus on monitoring for clinical illness, through nurse led symptom checks.

**Table I.**
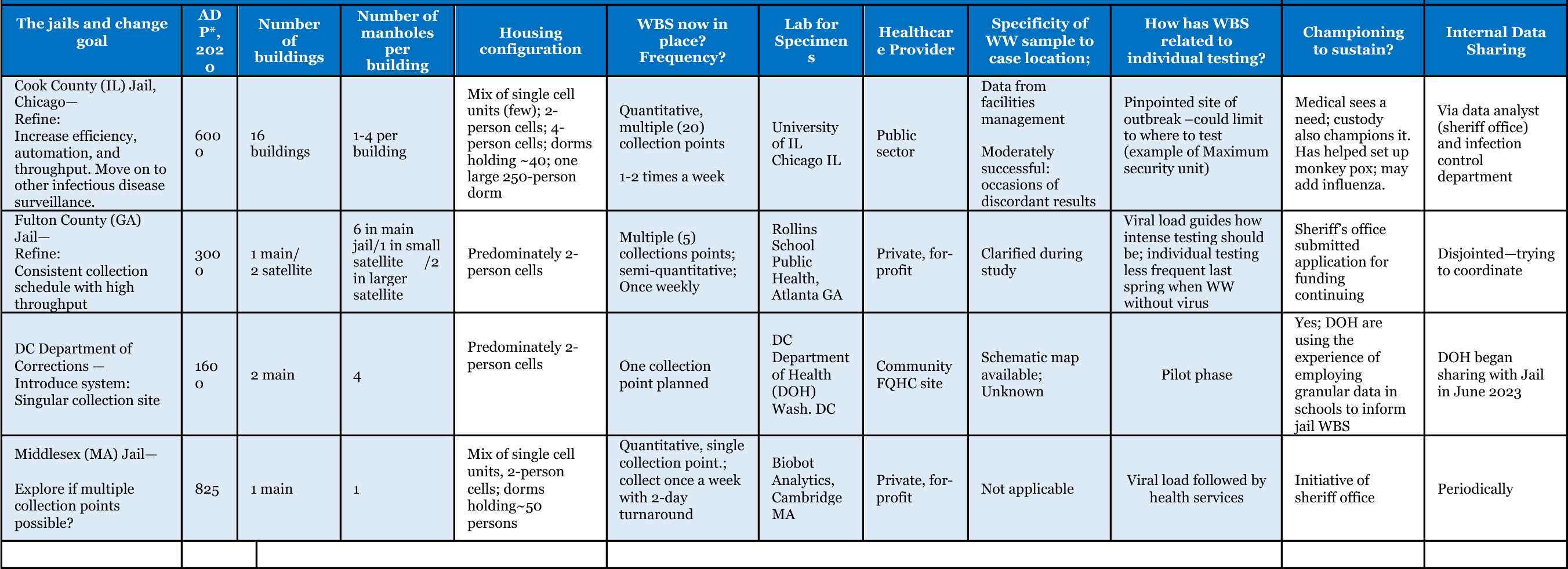
Preliminary Characteristics of Wastewater-based Surveillance (WBS) Methods among enrolled Jails (November 2022)

### A. Granularity of Wastewater Collection Sites at Cook County Jail and Fulton County Jail

At CCJ, each manhole access point drains from a single divisional living unit. This permitted infection control teams to plan for optimal isolation housing configurations and identify zones that did not require targeted testing. In October 2021, SARS-CoV-2 was detected in the wastewater from the maximum-security living unit after prolonged negative results. At the time, no persons were under investigation for infection. The infection control team used the positive wastewater readings as a harbinger of occult infection in this unit. They preemptively notified custody staff that new isolation beds were necessary. Transfers out of the building were stopped to prevent introducing new cases without proactive testing. Three days after the WBS signal appeared, clinical cases were diagnosed in the building. This early warning spared staff from scrambling for isolation beds and facilitated the prompt application of infection control measures.

Since the summer of 2021, wastewater results have been shared weekly with healthcare leadership at FCJ. For the entire duration of WBS, the jail population had exceeded capacity, limiting the ability to isolate and quarantine infected and exposed populations. Nonetheless, WBS spurred testing and led to isolation and quarantine as space permitted. Approaches were different during 2 periods:

<u>Period 1.</u> Summer 2021-Summer 2022: In the early summer of 2021, wastewater was clear of SARS-CoV-2 for several weeks. Healthcare staff identified no active cases. Later, the wastewater tested positive, preceding the detection of cases in the population. Because WBS could not pinpoint which housing units had cases, mass testing of hundreds of persons occurred beginning October 2021. This supplemented ongoing, opt-out antigen testing of entrants and symptomatic individuals in the population. Throughout 2022, the team increasingly understood the source of cases leading to positive wastewater sites (Supplement).

<u>Period 2.</u> Fall 2022: Locating the jail’s blueprints (Supplement and Textbox 1) permitted the Emory team to map plumbing lines. There are 42 housing units in the north tower, 36 in the south tower. The sewer mapping helped the Emory team narrow down the source site to 7 housing units (or 14 in the case where two spokes of a tower drained into one site). For example, in the week of October 17, wastewater was positive in the North 5 manhole but negative at North 6, prompting screening in levels 500 and 600 of the North Tower. A cluster of 4 cases was found in 2 North 500 and 0 cases in the latter.

### A. When Granular Results Are Not Necessary

MHOCJ had only one collection site and so case locations could not be pinpointed. Since infection could exist anywhere, many housing units were screened. Nonetheless, screening an entire jail of 825 persons was feasible, unlike a jail several magnitudes larger. In August 2021, a quick, high spike in wastewater counts coincided with the emergence of the Delta strain of COVID in MA (Figure 4b).

**Figure 4.**
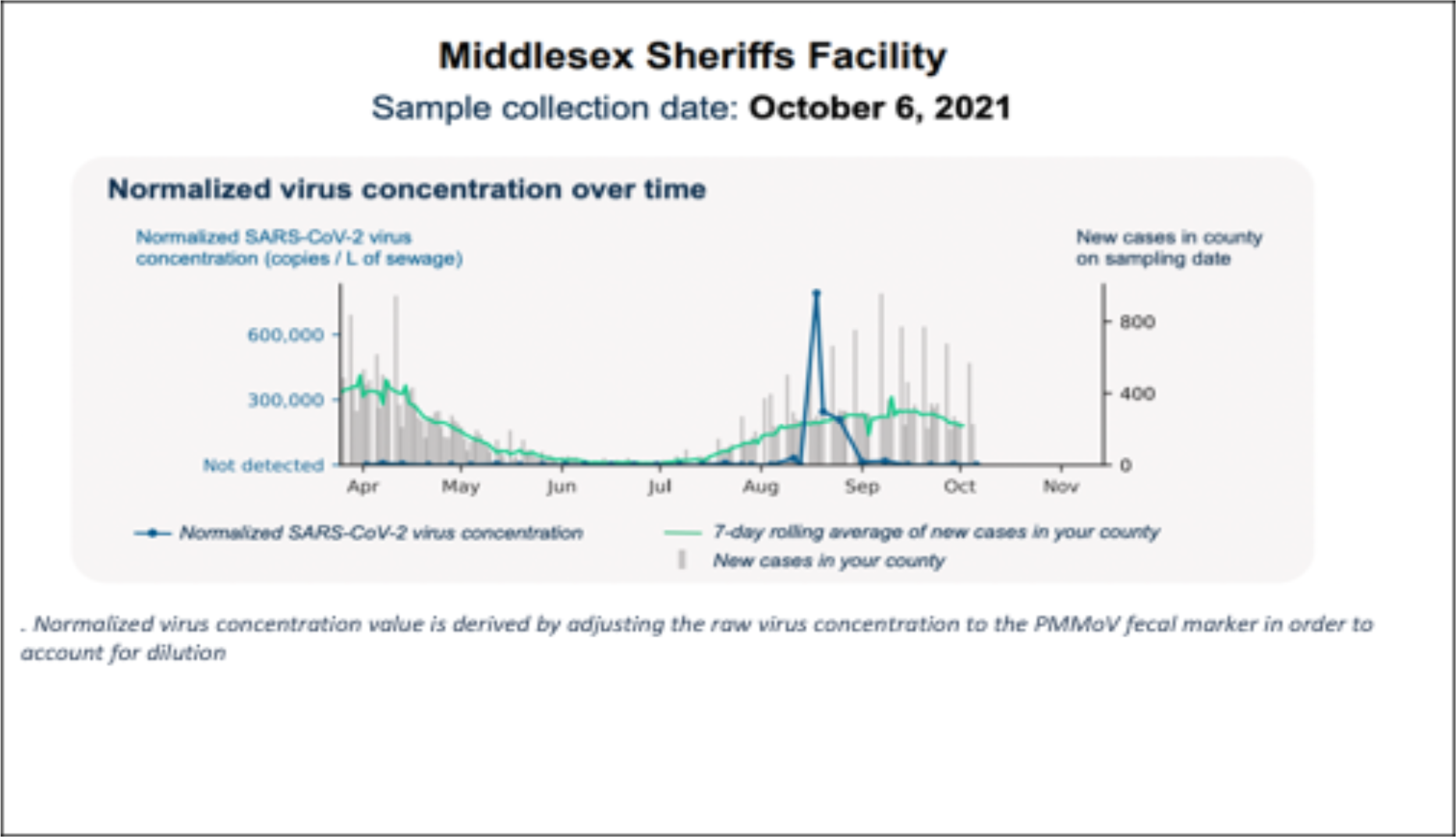
Graph of SARS-CoV-2 Virus concentration over time and new county cases in Middlesex County April 2021-October 2021.

### A. Improving WBS and Individual Collection Procedures

Under the aegis of the Emory team, future wastewater at FCJ will be collected using an automatic compiler, which is being installed at the time of this manuscript’s submission. The advantages of this technology are more regulated, periodic sampling of the wastewater, removing the need to drop Moore Swabs on one day and retrieve them the next. This is the same technology used at MHOCJ and the DCJ. When WBS indicates infection, several strategies, while respecting residents’ rights, can enhance case-finding efficiency. Successful tactics at FCJ, before testing could precisely locate the source of the virus in the wastewater, are included in the supplement.

## Conclusion

In the first year of WBS for SARS-CoV-2 in jails around the country, we learned that jails of varying sizes found WBS effective in monitoring new outbreaks. We describe four jail systems that now conduct surveillance by collecting WBS specimens and use the outcomes to mitigate outbreaks. Some were early adopters; issues with funding delayed the DC roll out. Each of the programs described here had unique, innovative approaches to WBS; however, they also had challenges to overcome.

WBS can indicate the emergence of infection after a period of no activity and prompt the swab testing of individuals. In larger jails, WBS may be more beneficial for knowing where to refrain from testing when cases are highly clustered. Each jail was half the size of the one ranked next largest. When a jail featured multiple collection points for wastewater in a large complex, we observed a lead time on case identification which allowed for advanced preparation of mitigation practices. These results streamlined resource distribution by indicating the areas of low viral load and, thus, areas that did not require advance preparation of quarantine or isolation beds. When a jail did not feature numerous wastewater collection points, or it was difficult to determine how the wastewater flow corresponded to specific housing units, testing needed to be widespread. FCJ demonstrated numerous nasal swabs could be collected quickly using barcode scanning to register specimens. Moreover, it also demonstrated the usefulness of dye testing to confirm wastewater flow. We believe that combining automatic compilation and high collection site granularity will be the most favorable, future practice. Lastly, MHOCJ shows that even a small jail can benefit from WBS technology.

Aside from jail size, the study team discovered that each site’s architectural features had implications for WBS. Configurations of jails varied from a collection of independent buildings, towers with multiple wings, or a singular structure. Each configuration posed different challenges. Residents were housed in dorms, double occupancy cells, single occupancy cells, or experienced mixed housing. The variance in housing was important to consider when planning individual testing and other mitigation strategies yet it demonstrated that WBS is a flexible model that can be applied to various configurations successfully.

The fundamental component underlying successful implementation of WBS in jails is support from various stakeholders. Across the country, early attitudes toward finding cases varied. They ranged from withholding testing of asymptomatic persons, even with known exposure,^31^ to aggressive identification of all cases and transparently posting results.^32^ Cooperation between entities included inter-jail cooperation and communication between custody officials and respective medical operations. FCJ and MHOCJ operated with contracted medical vendors who approved surveillance activity. External jail support included close relationships between jail staff and academic institutions like the University of Illinois in Chicago, Emory University, Montefiore Medical Center/Albert Einstein College of Medicine, and Tufts University. Funding entities made study activities possible as WBS is not yet considered standard population-level surveillance practice. Most importantly, jail residents were crucial to implementing the surveillance project, as their participation was necessary for calibrating individual testing logistics. The diversity represented in this study’s stakeholders was key to its success. Increased participation and awareness between different entities strengthened the process overall.

The results of this study suggest WBS could be a useful population-level surveillance system for other emerging infectious diseases such as Mpox, influenza, RSV, hepatitis A, polio, and tuberculosis. Wastewater was archived at CCJ and FCJ starting in May of 2022 at the onset of the Mpox outbreak. Validation of Mpox detection in wastewater has since been demonstrated.^33^ CCJ observed two confirmed cases of Mpox,^34^ but did not detect the virus in the archived, concurrent wastewater samples.

WBS opens the possibility of surveillance for non-infectious agents, such as opioids and other illicit drugs. Detection of substances in a sample representing the aggregate of the jail wastewater can provide public health data on what substances are present. Determining the precise location to narrow the search to a single housing unit could lead to an individual’s entrapment. The opinion of authors is divided: some support such a move, others believe it may erode residents’ trust in WBS programs. While targeted searches could save resources and better prevent overdose-associated morbidity and mortality, it could lead to associating WBS with punishment rather than health promotion.

We have demonstrated that WBS can serve as an early warning system for disease detection in carceral settings. Its potential to assist corrections and public health agencies with outbreak mitigation is enormous. The application needs to be thoughtful, and input from a wide range of stakeholders, including those with lived experience of incarceration could be useful when deciding its scope. Opinions may vary on whether to test wastewater for illicit substances. With all voices at the table, as its implementation is planned, the new technology could change the landscape of infection control in carceral settings and, by extension, other congregate environments like dormitories and homeless shelters.

## Supporting information

Title Page

## Data Availability

All data produced in the present study are available upon reasonable request to the authors

