## Supplementary figures and images for "Four Models of Wastewater-Based Surveillance for SARS-CoV-2 in Jail Settings: How Monitoring Wastewater Complements Individual Screening"

### Supplement

Supplement

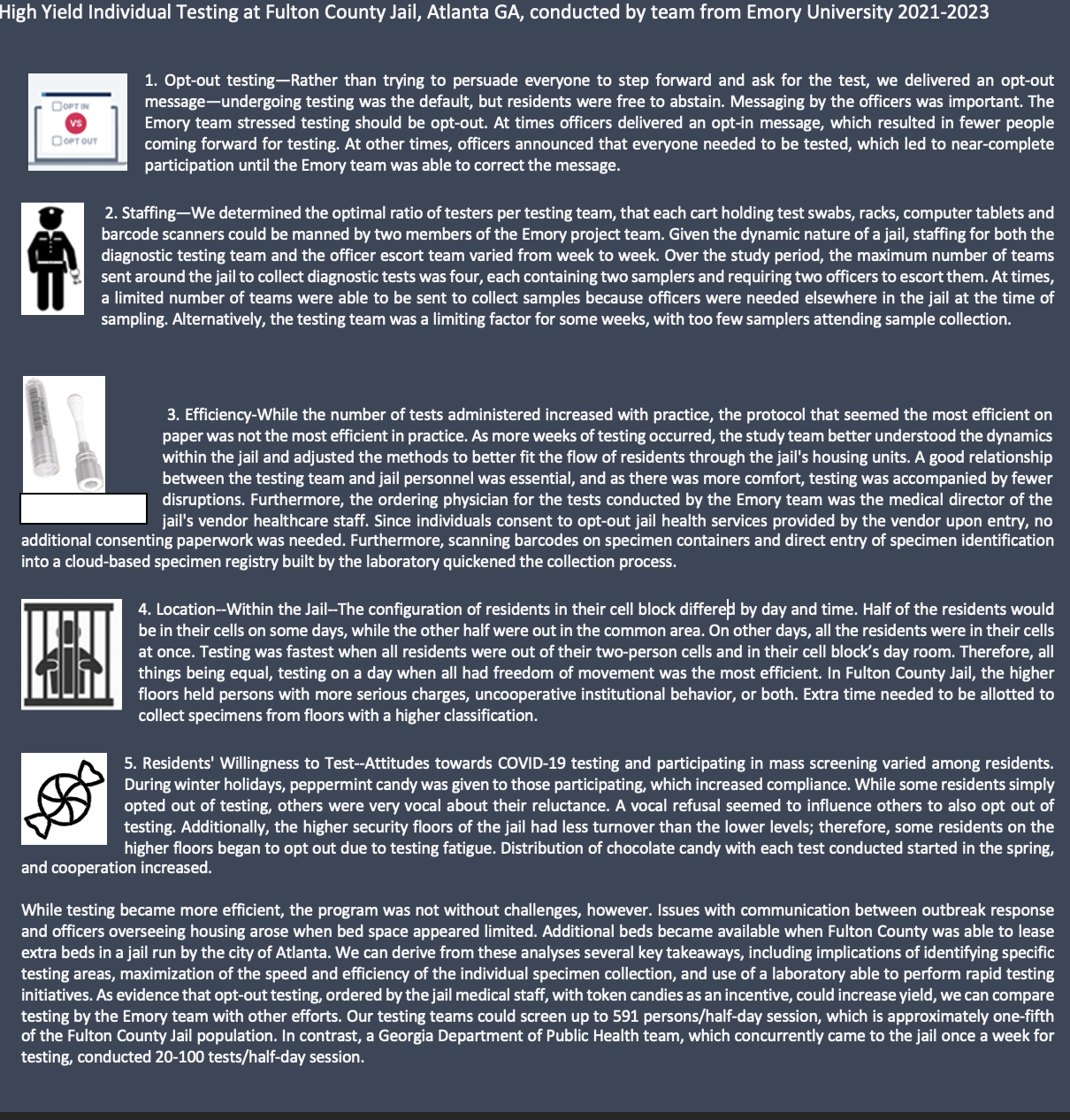
